# Pretreatment of patients with ST-segment elevation myocardial infarction with heparin: a systematic review and meta-analysis

**DOI:** 10.1101/2024.01.22.24301634

**Authors:** Gonçalo Costa, Bernardo Resende, Bárbara Oliveiros, Lino Gonçalves, Rogério Teixeira

## Abstract

**Background:** Unfractionated heparin (UFH) is frequently administered before percutaneous coronary intervention (PCI) in patients with ST-segment elevation myocardial infarction (STEMI). However, current guidelines do not provide clear recommendations for UFH pretreatment before arrival at the coronary catheterisation laboratory.

**Methods:** Between June and July 2023, we systematically searched PubMed, Embase and Cochrane databases for studies comparing UFH pretreatments in patients with STEMI. A random-effects meta-analysis and meta-regression analyses were performed.

**Results:** Fourteen studies were included, of which four were randomised clinical trials (RCTs). A total of 76446 patients were included: 31238 in the pretreatment group and 39208 in the control group. Our meta-analysis revealed a lower all-cause mortality for the pretreatment strategy when compared with the control group, albeit with high heterogeneity (pooled odds ratio (OR) = 0.61, 95% confidence interval (CI) [0.49 - 0.76], *P* < 0.01; I² = 77%); lower in-hospital cardiogenic shock (pooled OR = 0.68, 95% CI [0.58, 0.78], *P* < 0.21; I² = 27%) and a higher rate of spontaneous reperfusion events (pooled OR = 1.68, 95% CI [1.47, 1.91], *P* < 0.01; I² = 79%). In terms of major bleeding, the UFH pretreatment strategy further revealed a decreased rate of events (pooled OR = 0.85, 95% CI [0.73, 0.99], *P* = 0.40; I² = 4%).

**Conclusions:** Our study suggests that UFH pretreatment in patients with STEMI undergoing primary PCI was associated to reduced all-cause mortality, cardiogenic shock, enhancing reperfusion rates, whilst diminishing major bleeding events.

## INTRODUCTION

Acute ST-elevation myocardial infarction (STEMI) is a life-threatening condition that requires prompt reperfusion therapy to minimise myocardial damage and improve patient outcomes. Percutaneous coronary intervention (PCI) is widely recognised as the gold standard treatment for patient with STEMI as it helps restore coronary blood flow and salvage ischaemic myocardium (1). However, the optimal antithrombotic strategy for the pretreatment of patients with STEMI undergoing PCI remains a topic of debate.

One common approach is heparin administration prior to patient arrival at the coronary catheterisation laboratory. Heparin, specifically unfractionated heparin (UFH), has been previously used in clinical practice to improve spontaneous reperfusion rates and reduce the clot burden (2). Improved coronary blood flow prior to PCI, which positively impacts patient outcomes, has been previously demonstrated (3). UFH is a rapidly acting anticoagulant with a short half-life of approximately 1–2 hours after intravenous administration (4). Its pharmacokinetic profile, coupled with antidote availability, means that UFH is a potential early-administration candidate in patients with STEMI.

Despite the widespread use of UFH during PCI, robust evidence is scarce on the benefits of administering anticoagulation at earlier stages within a PCI context. To date, only four small randomised controlled trials (RCTs) (2,5–7) and several observational studies (8–17) with varying results on patient-relevant outcomes have investigated UFH pretreatment in patients with STEMI undergoing PCI. Additionally, the evidence on mortality outcomes remains inconclusive. Consequently, recent European guidelines on acute coronary syndromes have endorsed UFH during PCI but they do not provide clear recommendations for UFH pretreatment before arrival at the coronary catheterisation laboratory (1).

Therefore, we conducted a systematic review and meta-analysis to evaluate efficacy and safety outcomes associated with UFH pretreatment in patients with STEMI undergoing primary PCI.

## METHODS

### Protocol and Registration

This study was designed according to the Preferred Reporting Items for Systematic Reviews and Meta-Analyses statement (**Supplementary Table 1**). This systematic review and meta-analysis were registered with the PROSPERO database (CRD42023422529).

In total, we made two changes to the original registry. Firstly, we expanded the scope of secondary outcomes to encompass stroke and major adverse cardiovascular outcomes. Additionally, we excluded post-PCI Thrombolysis in Myocardial Infarction (TIMI) flow-grade and no-reflow phenomena due to insufficient data. Secondly, we substituted the control arm, replacing UFH administration at the catheterisation laboratory with either no UFH pretreatment or delayed administration. This alteration mitigated a potential major limitation in study selection.

### Literature Searches

We systematically checked the Cochrane Controlled Register of Trials (CENTRAL), EMBASE and PubMed between June and July 2023. In all databases, we accessed both interventional and observational studies which compared heparin pretreatment with delayed heparin administration in patients with STEMI in multiple combinations. Our selection criteria had no language or date restrictions. We also analysed bibliographic references in eligible studies to capture additional articles via cross referencing. The meta-analysis search and selection strategy is shown (**Figure 1**).

**Figure 1.**
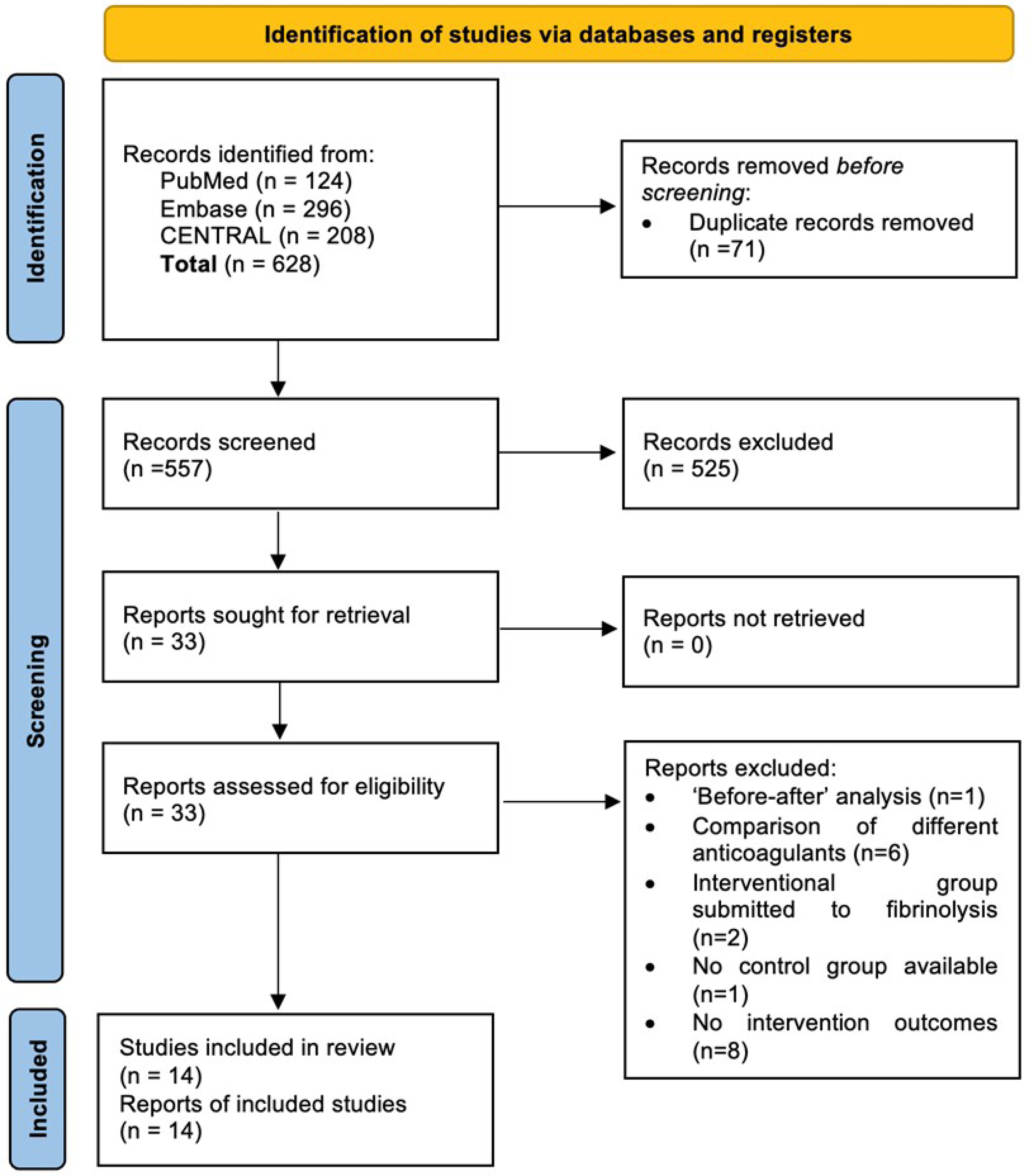
Literature search flow diagram.

### Eligibility Criteria

We used the following criteria to define study eligibility: 1) studies comparing the use of UFH pretreatment, prior to arrival at the catheterisation laboratory, with either no UFH pretreatment or delayed administration and 2) studies describing UFH administration timing and doses. We excluded studies reporting the use of other anticoagulant types as pretreatments that did not encompass full-text article publications and without control groups.

### Primary and Secondary Outcomes

Primary outcomes were all-cause mortality and major bleeding events. Secondary endpoints were in-hospital cardiogenic shock, spontaneous reperfusion (defined as pre-PCI TIMI flow 2–3), stroke, cardiovascular mortality, major adverse cardiovascular outcomes, minor bleeding events and all bleeding events.

### Data Collection and Management

Two authors systematically reviewed titles and abstracts of retrieved publications to identify studies meeting inclusion criteria. Any disagreements on study eligibility were resolved by consensus and discussion. Data collected from selected studies were subject to a narrative synthesis approach covering the study population, including key demographics and clinical characteristics, interventions, and outcomes. For studies with multiple sequential publications, measures were taken to avoid the duplication of results. When summary data were not readily available, calculations were performed using available study data.

### Risk of Bias and Certainty Assessments

Two authors independently assessed the risk of bias in articles according to the Cochrane Collaboration’s “Risk of bias” tool for RCTs and the Newcastle–Ottawa Scale for observational studies. Studies with a high risk of bias were excluded. Quality evaluations for studies are presented in the “risk of bias summary” table (**Table 1**) or in the Newcastle–Ottawa Scale summary table (**Table 2**). Regarding certainty of evidence we used Grading of Recommendations, Assessment, Development and Evaluations framework.

**Table 1.** Risk of bias summary. Green circle – low risk of bias; Red circle – high circle of bias.

|  | Random sequence generation (selection bias) | Allocation concealment (selection bias) | Blinding of participants and personnel (performance bias) | Blinding of outcome assessment (detection bias) | Incomplete outcome data (attrition bias) | Selective reporting (reporting bias) | Other bias |
| --- | --- | --- | --- | --- | --- | --- | --- |
| Braga et al 1998 | + |  | + |  | + | + | + |
| Cantor et al 2017 | + | + | - | + | + | + | + |
| Fakhr-Mousavi et al 2021 | + |  | - | + | - | - | + |
| Karlsson et al 2019 | + | + | - | - | + | + | + |

**Table 2.** Newcastle–Ottawa Scale summary. * - 1 point.

| Study | Selection | Comparability | Outcome | Total |
| --- | --- | --- | --- | --- |
| Ariza et al, 2013 | **** | - | * | 5 |
| Bloom et al, 2021 | **** | - | *** | 7 |
| d'Entremont et al, 2020 | *** | ** | * | 6 |
| Emilsson et al, 2022 | **** | ** | *** | 9 |
| Giralt et al, 2015 | **** | ** | *** | 9 |
| Giralt et al, 2020 | **** | - | *** | 7 |
| McGinley et al, 2020 | *** | ** | *** | 8 |
| Verheugt et al, 1998 | *** | * | *** | 7 |
| Żurowska-Wolak et al, 2021 | **** | - | *** | 7 |

### Statistical Analysis

We conducted our meta-analysis to pool data across studies using a random effects model. The mean effect was considered significant if its 95% confidence interval (CI) did not include zero. Outcomes were addressed by estimating odds ratios (ORs). Statistical heterogeneity was evaluated by the visual inspection of forest plots, the I² statistical index (< 25% low, 25%–40% moderate and > 40% high heterogeneity) and Egger’s linear regression tests. The software package used for the meta-analysis was Review Manager (RevMan) version 5.4.1. Meta-regression was performed considering one independent variable each time to generate regression model assumptions, such as the absence of multicollinearity and at least 5–10 cases (case studies) per independent variable. Moreover, due to limited complete pairwise data, multiple meta-regression was not valid. We applied a random-effects model using DerSimonian–Laird and Knapp–Hartung standard error adjustment methods using meta-regression procedures in IBM SPSS, version 28. All data were evaluated at a 5% significance level (P<0.05).

## RESULTS

### Search Results

Our literature search identified 628 relevant records. Following duplicate removal, 525 publications were excluded based on pre-defined inclusion and exclusion criteria. Finally, 14 studies, 4 RCTs (2,5–7) and 10 observational studies (8–17), were included **(Figure 1)** with a total of 76446 patients: 31238 in the pretreatment group and 39208 in the control group. Most studies used UFH doses between 5000-10000 U or 90 U/kg. Two studies opted for a high dose of 300 U/kg in the intervention arm. The timing of pretreatment varied, often given during transport to a PCI-capable hospital. While most studies mention aspirin loading, the choice of P2Y12 inhibitors varied, with clopidogrel being the most common. Study characteristics of selected studies are presented in **Table 3** and baseline patient characteristics are summarised in **Table 4**.

**Table 3.** *Characteristics of selected studies.* ACT = Activated Clotting Time, CL = Catheterisation Laboratory, ER = Emergency Room, GpIIb/IIIa = Glycoprotein IIb/IIIa, IV = Intravenous, NR = Not Reported, PCI = Percutaneous Coronary Intervention, SC = Subcutaneous, UF = Unfractionated Heparin.

| Study | Design | n (%) |  | Timing of UFH administration |  | IV UFH dose scheme |  | Concomitant antiplatelet drug: Dose, n (%) |  | Follow-up |
| --- | --- | --- | --- | --- | --- | --- | --- | --- | --- | --- |
|  |  | Pretreatment group | Control group | Pretreatment group | Control group | Pretreatment group | Control group | Pretreatment group | Control group |  |
| Ariza et al, 2013 | Prospective | 566 (57.8) | 414 (42.2) | Transport or ER | CL | 0.75-1mg/Kg and extra dose cording to ACT | 1 mg/Kg (8% of the patients received bivalirudin) | • Aspirin, NR: 566 (100.0)<br>• Clopidogrel: 600mg, 538 (95.0) | • Aspirin: NR, 412 (99.5)<br>• Clopidogrel: 300 or 600mg, 401 (98.1) | Evaluation pre and post PCI |
| Bloom et al, 2021 | Retrospective | 1392(29.5) | 3328(70.5) | Transport | ER or CL | 4000U plus 1000U in 1 hour | NR | • Aspirin: NR, 1346 (96.7)<br>• Ticagrelor: NR, 947 (68.0) | • Aspirin: NR 3148 (94.6)<br>• Ticagrelor: NR, 2140 (64.3)<br>• GpIIb/IIIa inhibitor: NR, 1281 (38.5) | 30 days |
| Braga et al, 1998 | Randomized clinical trial | 25 (52.1) | 23 (47.9) | ER | CL | 300U/Kg | 5000-10000U | • Aspirin: 200mg, 25 (100.0) | • Aspirin: 200mg, 23 (100.0) | Evaluation pre and post PCI |
| Cantor et al, 2019 | Randomized clinical trial subanalysis | 5422 (65.2) | 2,889 (34.8) | Transport or ER | CL | 7,688.3 ± 3,686.9U | 6,936.6 ± 3,358.9U | • Aspirin: NR, NR<br>• Clopidogrel: NR, NR<br>• Prasugrel: NR, NR<br>• Ticagrelor: NR, NR | • Aspirin: NR, NR<br>• Clopidogrel: NR, NR<br>• Prasugrel: NR, NR<br>• Ticagrelor: NR, NR | 1 year |
| Chung et al, 2007 | Retrospective | 56 (46.7) | 64 (53.3) | Transport or ER | After PCI | 60U/Kg plus 14U/Kg/h until PCI or 1mg/Kg SC enoxaparin | 100U/Kg | • Aspirin: 300mg, 56 (100.0)<br>• Clopidogrel: 300mg, 56 (100.0) | • Aspirin: 300mg, 64 (100.0)<br>• Clopidogrel: 300mg, 64 (100.0) | 8.6±5.0 days |
| d'Entremont et al, 2020 | Retrospective | 482 (68.0) | 227 (32.0) | Non-PCI capable center before transport | CL | 60U/Kg | 60U/Kg | • Aspirin: 320mg, 482 (100.0)<br>• Ticagrelor: 180mg, 482 (100.0) | • Aspirin: 320mg, 227 (100.0)<br>• Ticagrelor: 180mg, 227 (100.0) | NR |
| Emilsson et al, 2022 | Retrospective | 16026 (38.5) | 25605 (61.5) | NR | CL | NR | NR | • Aspirin: NR, 15332 (95.7)<br>• Clopidogrel: NR, 5581 (34.8)<br>• Ticagrelor: NR, 7688 (48.0) | • Aspirin: NR, 25112 (98.1)<br>• Clopidogrel: NR, 14986 (58.5)<br>• Ticagrelor: NR, 10120 (39.6) | 30 days |
| Fakhr-Mousavi et al, 2021 | Randomized clinical trial | 92 (54.4) | 77 (45.6) | ER | CL | 90U/Kg | 90U/kg | • Aspirin: 325mg, 92 (100.0)<br>• Clopidogrel: 600mg, 92 (100.0) | • Aspirin: 325mg, 77 (100.0)<br>• Clopidogrel: 600mg, 77 (100.0) | 40 days |
| Giralt et al, 2015 | Retrospective | 758 (57.2) | 568 (42.8) | Non-PCI capable center before transport or during transport | CL | 5000U | 5000U | • Aspirin: 300mg, 744 (98.2)<br>• Clopidogrel: 600mg, 742 (97.9) | • Aspirin: 300mg, 531 (93.5)<br>• Clopidogrel: 300mg, 503 (88.6) | 1 year |
| Giralt et al, 2020 | Prospective | 2720 (77.3) | 800 (22.7) | >31 minutes before PCI | Administered <30 minutes before PCI | 70-100 U/Kg (5000U maximum) | 70-100 U/Kg (5000U maximum) | • Clopidogrel: 600mg, 1973 (72.5)<br>• Ticagrelor or Prasugrel:180 or 60mg, 654 (24.0) | • Aspirin: 250-300mg, 655 (81.9)<br>• Clopidogrel: 600mg, 492 (61.5)<br>• Ticagrelor or Prasugrel:180 or 60mg, 117 (14.6) | 1 year |
| Karlsson et al, 2019 | Randomized clinical trial subanalysis | 2898 (40.6) | 4246 (59.4) | Non-PCI capable center before transport or during transport or in ER | CL | NR | NR | • Aspirin: NR, 2817 (97.2)<br>• Clopidogrel: NR, 1662 (57.4)<br>• Ticagrelor: NR, 731 (25.2) | • Aspirin: NR, 3636 (85.6)<br>• Clopidogrel: NR, 2489 (58.6)<br>• Ticagrelor: NR, 578 (13.6) | 30 days |
| McGinley et al, 2020 | Retrospective | 437 (43.7) | 563 (56.3) | In transport | CL | 5000U | NR | • Aspirin: 300mg, 437 (100.0)<br>• Clopidogrel: 300mg, 437 (100.0) | NR | 5 years |
| Verheugt et al, 1998 | Quasi-experimental comparative study | 108 (50.0) | 108 (50.0) | ER | CL | 300U/Kg | NR | • Aspirin: 160mg, 108 (100.0) | • Aspirin: 160mg, 108 (100.0) | Evaluation pre and post PCI |
| Żurowska-Wolak et al, 2021 | Retrospective | 256 (46.4) | 296 (53.6) | Transport | ER or CL | NR | NR | • Clopidogrel: NR, NR | • Clopidogrel: NR, NR | 1 year |

**Table 4.** *Patient demographics and baseline characteristics.* NR = Not Reported, SD = Standard deviation.

| Study | Age<br>Mean±SD |  | Male sex<br>n (%) |  | Hypertension<br>n (%) |  | Diabetes mellitus<br>n (%) |  | Current smoking<br>n (%) |  | Door-to-balloon<br>time (min) |  | Killip class, I/II/III/IV<br>(%) |  |
| --- | --- | --- | --- | --- | --- | --- | --- | --- | --- | --- | --- | --- | --- | --- |
|  | Pretreatment<br>group | Control<br>group | Pretreatment<br>group | Control<br>group | Pretreatment<br>group | Control<br>group | Pretreatment<br>group | Control<br>group | Pretreatment<br>group | Control<br>group | Pretreatment<br>group | Control<br>group | Pretreatment<br>group | Control group |
| Ariza et al, 2013 | 61.6±12.4 | 62.2±13.8 | 450 (79.5) | 324 (78.3) | 301 (53.2) | 234 (56.5) | 133 (23.5) | 102 (24.6) | 272 (48.0) | 196 (47.3) | 73.2 ± 36.3 | 66.5 ± 37.1 | NR | NR |
| Bloom et al, 2021 | 62.0±12.4 | 64.0±13.2 | 1116 (80.2) | 2505 (75.3) | NR | NR | 203 (14.6) | 583 (17.5) | NR | NR | 48 ± 23 | 64 ± 47 | NR | NR |
| Braga et al, 1998 | 62.0±12.0 | 64.0±10.0 | 16 (64.0) | 13 (57.0) | 13 (52.0) | 15 (65.0) | 5 (20.0) | 3 (13.0) | 16 (64.0) | 11 (48.0) | 71.0 ± 52.0 | 68.0 ± 17.0 | NR | NR |
| Cantor et al, 2019 | 61.1±11.8 | 60.9±12.0 | 4955 (77.7) | 2843 (77.2) | 3,194 (50.1) | 1,854 (50.4) | 1,208 (18.9) | 647 (17.6) | 2,908 (45.6) | 1,690 (45.9) | 86.6 ± 329.0 | 60.9 ± 108.2 | NR | NR |
| Chung et al, 2007 | 61.0±13.0 | 60.0±14.0 | 48 (85.7) | 52 (81.3) | 25 (45.0) | 37 (58.0) | 16 (29.0) | 14 (22.0) | 28 (50.0) | 25.0 (39.0) | 125±65 | 124±53 | 68.0/14.0/2.0/16.0 | 66.0/3.0/4.0/15.0 |
| d'Entremont et al, 2020 | 62.7±12.0 | 64.2±12.1 | 368 (76.3) | 174 (76.7) | 215 (44.7) | 114 (50.2) | 83 (17.3) | 32 (14.1) | 205 (42.6) | 118 (52.0) | 118 (98,146) | 87 (70, 100) | NR | NR |
| Emilsson et al, 2022 | 67.0±12.0 | 67.0±12.0 | 11259 (70.0) | 18,381 (71.8) | 7,118 (44.4) | 11,430 (44.6) | 2,318 (14.5) | 3,812 (14.9) | 4,662 (29.1) | 6,514 (25.4) | 276±244 | 290±270 | NR | NR |
| Fakhr-Mousavi et al, 2022 | 57.1±8.8 | 57.5±7.5 | 72.0 (78.3) | 60 (77.9) | 32 (34.8) | 41 (53.2) | 13 (14.1) | 23 (29.9) | NR | NR | NR | NR | 96.7/2.2/1.1/- | 97.4/2.6/-/- |
| Giralt et al, 2015 | 61.3±12.8 | 63.4±12.8 | 618 (81.5) | 434 (76.4) | 407 (53.7) | 291 (51.2) | 163 (21.5) | 140 (24.6) | 344 (45.4) | 237 (41.7) | 107 (86,133) | 105 (83,140) | 85.0/9.1/2.0/4.0 | 74.1/13.4/4.9/7.6 |
| Giralt et al, 2020 | 65.4±14.0 | • 994 patients: 62.6±12.8<br>• 1091 patients: 62.3±12.8<br>• 635 patients: 64.4±13.4 | 2173 (79.9) | 600 (75.0) | 1417 (52.1) | 461 (57.6) | 464 (17.1) | 150 (18.8) | 1190 (43.8) | 281 (35.1) | • 994 patients: 82.0 (68.0;103)<br>• 1091 patients: 101 (88.0;122)<br>• 635 patients: 140 (121;175) | 109 (78.0,161) | 82.5/8.0/2.2/5.9 | 79.8/7.4/2.8/10.0 |
| Karlsson et al, 2019 | 66.0±11.5 | 66.0±11.6 | 2169 (74.8) | 3177 (74.8) | 1172 (40.8) | 1850 (44.3) | 373 (12.9) | 514 (12.2) | 987 (35.0) | 1245 (31.9) | 185 (125, 320) | 181 (118, 327) | NR | NR |
| McGinley et al, 2020 | 63.7±NR | 63.7±NR | 304 (69.6) | 390 (69.3) | 132 (30.2) | 168 (29.8) | 38 (8.7) | 53 (9.4) | 173 (39.6) | 232 (41.2) | NR | NR | NR | NR |
| Verheugt et al, 1998 | NR | NR | 91 (84.0) | 91 (84.0) | NR | NR | NR | NR | NR | NR | 132 (18, 360) | 132 (48, 360) | NR | NR |
| Zurowska-Wolak et al, 2021 | 64.4±11.8 |  | NR | NR | 384 (69.6) |  | 144 (26.1) |  | NR | NR | 87 ±29 |  | NR | NR |

### Primary Outcomes

Our meta-analysis identified lower all-cause mortality rates using a pretreatment strategy when compared with a control group, albeit with high heterogeneity (pooled OR = 0.61, 95% CI [0.49, 0.76], *P* < 0.01; I² = 77%). In subgroup analyses, in-hospital and 30-day mortality maintained statistical significance (pooled OR = 0.60, 95% CI [0.42, 0.86], *P* < 0.41; I² = 0%; pooled OR = 0.63, 95% CI [0.46, 0.86], *P* < 0.01; I² = 81%, respectively). A reduction in mortality effects was also observed for 1-year all-cause mortality, but with high heterogeneity (pooled OR = 0.56, 95% CI [0.32, 0.96], *P* < 0.01; I² = 91%) (**Figure 2, Figure S1**). In meta-regression analyses, age ratio (b = - 0.90; p = 0.786), aspirin ratio (b = -0.81; p = 0.378), PY12 inhibitor ratio (b = -0.49; p = 0.351), ticagrelor ratio (b = -0.06; p = 0.519) or ‘time to PCI’ (b = 0.18; p = 0.519) were not predictors of the evaluated effect size. However, the effect size tended to be greater when the clopidogrel ratio was smaller (b = -0.89; p = 0.026) (**Figure S2**).

**Figure 2.**
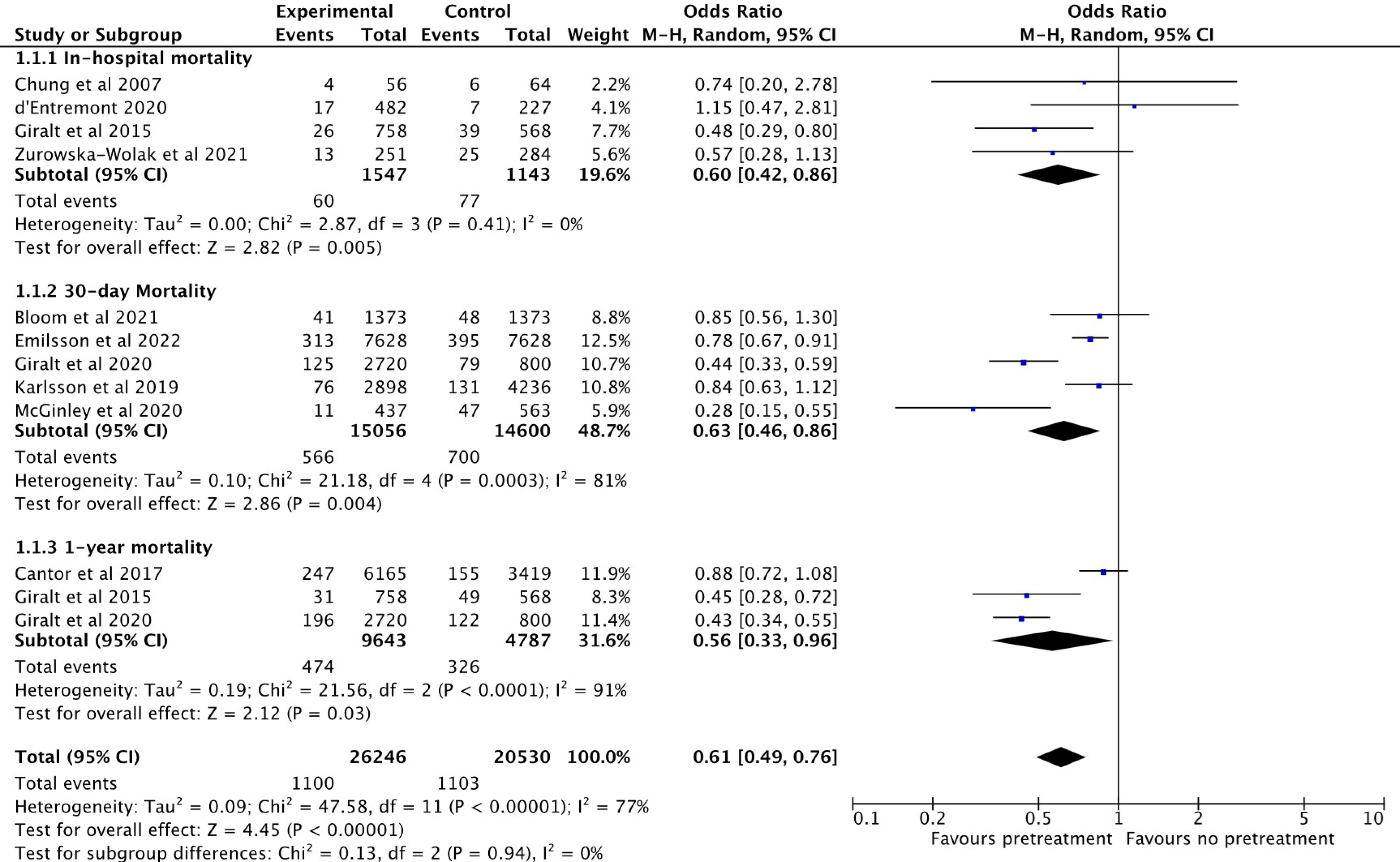
Forest plot showing all-cause mortality comparing unfractionated heparin pretreatment versus delayed administration. M–H, Mantel–Haenszel.

In terms of major bleeding, the pretreatment strategy further revealed a decreased rate of events (pooled OR = 0.85, 95% CI [0.73, 0.99], *P* = 0.40; I² = 4%) (**Figure 3, Figure S3**).

**Figure 3.**
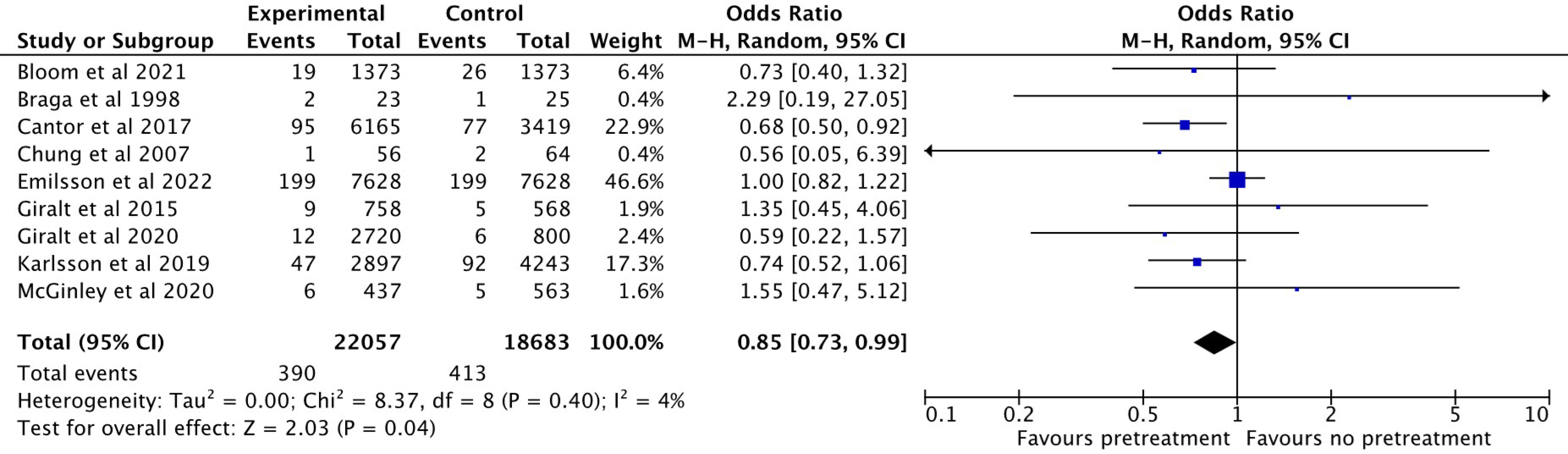
Forest plot showing major bleeding events comparing unfractionated heparin pretreatment versus delayed administration. M–H, Mantel–Haenszel.

### Secondary Outcomes

In terms of secondary outcomes, in-hospital cardiogenic shock was statistically significantly favourable for the pretreatment strategy (pooled OR = 0.68, 95% CI [0.58, 0.78], *P* < 0.21; I² = 27%) (**Figure 4, Figure S4**). Additionally, the pretreatment strategy exhibited a significantly higher rate of spontaneous reperfusion events (pooled OR = 1.68, 95% CI [1.47, 1.91], *P* < 0.01; I² = 79%) (**Figure 5, Figure S5**) and stroke (pooled OR = 0.94, 95% CI [0.66, 1.34], *P* = 0.98; I² = 0%). Cardiovascular mortality, major adverse cardiovascular events, minor bleeding and all-bleeding analyses were not performed due to insufficient data reported in the included studies.

**Figure 4.**
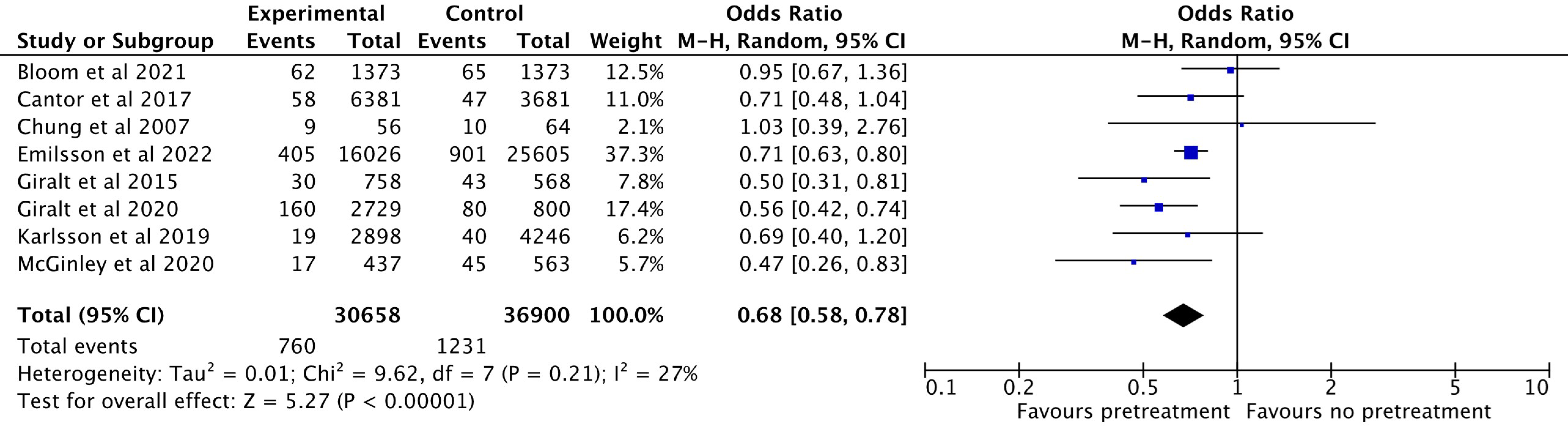
Forest plot showing in-hospital cardiogenic shock comparing unfractionated heparin pretreatment versus delayed administration. M–H, Mantel–Haenszel.

**Figure 5.**
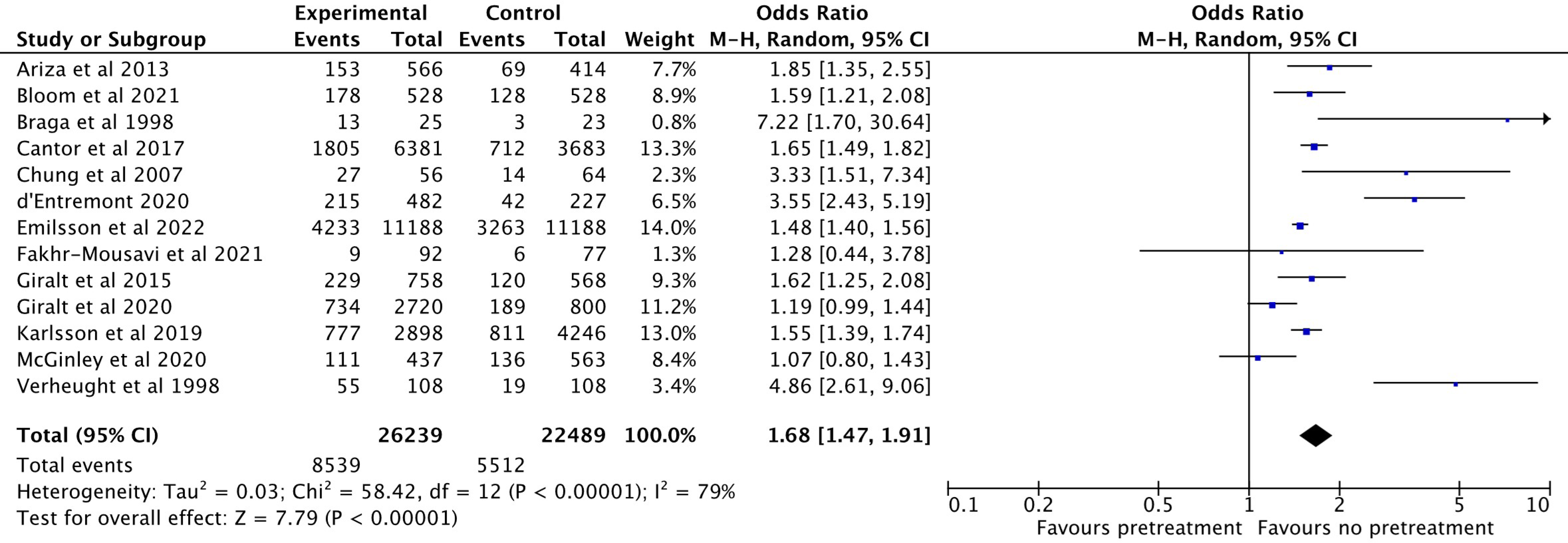
Forest plot showing spontaneous reperfusion events (pre-percutaneous coronary intervention Thrombolysis in Myocardial Infarction flow 2–3) comparing unfractionated heparin pretreatment versus delayed administration. M–H, Mantel– Haensz

### Risk of Bias and Evidence Certainty

Overall, selected studies demonstrated a low–moderate risk of bias. However, due to the nature of the intervention, a high risk of bias was observed in participant and personnel blinding (**Tables 3 and 4**). Therefore, considering our results and the risk of bias in terms of our robust evidence, we considered low certainty for primary outcomes and very low for secondary outcomes (**Supplementary Table 2**).

## DISCUSSION

We performed a systematic review and meta-analysis on the efficacy and safety outcomes of UFH pretreatment in patients with STEMI undergoing primary PCI. We observed that UFH pretreatment was associated with a lower risk of all-cause mortality, in-hospital cardiogenic shock and spontaneous reperfusion events. Furthermore, an improvement in safety outcomes was observed for reduced major bleeding events between groups.

Current guidelines recommend adjunctive antithrombotic treatment with antiplatelet and anticoagulant medication before primary PCI in patients diagnosed with STEMI, but ideal administration times remain controversial (1).

The notable reduction in all-cause mortality associated with UFH pretreatment underscores its potential as a valuable intervention; enhanced coronary blood flow before primary PCI which mitigates myocardial damage is compelling. Previous studies have suggested that UFH, as a rapidly acting anticoagulant, may facilitate spontaneous reperfusion rates and reduce the clot burden (2). This mechanism is particularly relevant within the STEMI context, where rapid and effective reperfusion is a critical determinant of patient outcomes (3), as shown by favourable outcomes in in-hospital, 30-day and 1-year mortality in our analyses. These findings further support the potential use of UFH pretreatment during critical post-PCI phases and in reducing early mortality.

By aligning the potential benefits of efficacy effects with safety considerations, the assessment of major bleeding events revealed a significant difference in the pretreatment group when compared with the control group. This observation did not exclude safety clinical evaluations before UFH pretreatment, especially given the delicate balance between preventing thrombotic events and avoiding bleeding complications (18). While acknowledging our results for this outcome, the caveat is that not enough data are available to support this conclusion when applied to elderly and frail patients.

Beyond mortality and haemorrhagic events, a substantial reduction in in-hospital cardiogenic shock associated with UFH pretreatment has profound clinical implications. Cardiogenic shock is a pivotal determinant of patient prognosis post-STEMI (19). Previous studies have demonstrated associations between successful reperfusion and a reduced risk of cardiogenic shock (20), so any intervention that potentially mitigates its occurrence warrants careful consideration. The observed reduction in this high-risk complication suggests that UFH pretreatment may help improve haemodynamic stability during acute STEMI phases, thus potentially impacting the broader course of the disease.

Moreover, our analyses identified a significant increase in spontaneous reperfusion event rates upon UFH pretreatment. Swift and effective reperfusion lies at the core of STEMI management (3), and this finding aligns with the mechanistic rationale underpinning UFH pretreatment. The potential for UFH to enhance the early restoration of coronary blood flow introduces an intriguing facet to the clinical benefits of the strategy. This prompts further exploration into the mechanisms underlying these outcomes, potentially involving thrombus burden reduction, and enhancing coronary flow.

In light of recent research, the study by Emilsson et al. (12), a well-conducted and -powered study, provides a valuable addition to the UFH pretreatment debate in patients with STEMI. By analysing data from the Swedish Coronary Angiography and Angioplasty Registry, Emilsson’s research aligns with our findings. The identification of a significantly lower risk of 30-day mortality, major bleeding events and cardiogenic shock lends further credence to the potential benefits of UFH pretreatment in improving patient outcomes.

Moreover, in two studies by Giralt et al (13,14), both having significant weight in our meta-analysis, the authors witnessed a sustained reduction in mortality outcomes, whether in-hospital, 30-day or 1-year mortality, and lower rates of in-hospital cardiogenic shock. However, both analyses showed similar grades of major bleeding events between groups despite a slight no-benefit-tendency in the pretreatment group. We observed that the mean door-to-balloon time in Emilson et al. was somewhat higher when compared with the studies by Giralt et al., which may explain these differences.

Collectively, our systematic review and meta-analysis provide crucial insights into the potential benefits and limitations of UFH pretreatment in primary PCI for patients with STEMI. The reduction in all-cause mortality, coupled with improvements in in-hospital cardiogenic shock and spontaneous reperfusion rates, underscores the potential advantages of this strategy. Although the absence of significant differences in major bleeding and stroke suggests potential safety issues, further exploration is warranted. Rigorous, large-scale RCTs with standardised protocols and reported outcomes are pivotal in establishing definitive efficacy and safety evidence for UFH pretreatment. This evidence will not only guide clinical decision-making but also optimise outcomes in the dynamic STEMI management field.

### Study Limitations

By acknowledging study strengths and limitations, some aspects of our research require careful consideration. Inherent heterogeneity across selected studies, stemming from variations in patient characteristics, UFH dosages and different concomitant antiplatelet regimens, may have introduced potential biases and limited the generalisability of our findings. A major difficulty in our study was different UFH administration timings in the pretreatment group, which limited the determination of exact administration times for anticoagulation and PCI treatments. Due to a lack of data in selected studies, we were unable to assess heparin drug-related problems such as heparin-induced thrombocytopenia or osteopenia (21,22). Due to the low incidence of these issues (23), we hypothesize that they exert a low impact on the choice of UFH as an anticoagulant therapy.

Despite these challenges, we addressed these concerns by analysing a significative number of studies using strong statistical methods and robust subgroup analyses. The absence of comprehensive reporting for specific outcomes, such as cardiovascular mortality, major adverse cardiovascular events, minor bleeding, and all bleeding events, underscores the importance of standardised reporting in future research.

## CONCLUSION

From our study, UFH pretreatment in patients with STEMI undergoing primary PCI has potential promise in reducing mortality, cardiogenic shock and enhancing reperfusion rates. High-scale RCTs are required to address these clinical questions in the future.

## Data Availability

All included studies are publicaly available

## ABREVIATIONS

CENTRAL: Cochrane Controlled Register of Trials
CI: confidence interval
ORs: odds ratios
PCI: percutaneous coronary intervention
RCTs: randomized clinical trials
SCAAR: Swedish Coronary Angiography and Angioplasty Registry
STEMI: ST-segment elevation myocardial infarction
TIMI: Thrombolysis in Myocardial Infarction
UFH: Unfractionated heparin

## Author Contributions

GC: Conceptualization, Data curation, Formal analysis, Methodology, Project administration, Writing – original draft; BR: Data curation, Formal analysis, Methodology, Writing – original draft; BO: Supervision, Formal analysis; LG: Supervision, Writing - review & editing; RT: conception and design, interpretation of data, revision of the manuscript, final approval and is responsible for the overall content as guarantor.

